# Emboli Transport in a Full-Length Patient-Specific Aorta: Assessment of Abdominal Organ Injury Risk During Cardiopulmonary Bypass

**DOI:** 10.1101/2025.06.07.25329120

**Authors:** Nafis M. Arefin, Bryan C. Good

**Author notes:** **Author for correspondence:** Bryan C. Good,.

## Abstract

Cardiopulmonary bypass (CPB), though indispensable in cardiac surgery, carries significant risks of systemic embolization and organ injury. While cerebral and cardiac complications have been thoroughly investigated, the impact of emboli on abdominal organs remains largely unexplored. This study integrates computational fluid dynamics (CFD) with Lagrangian particle tracking (LPT) to simulate emboli transport and hemodynamics under clinically relevant CPB conditions in a patient-specific aorta model. A validated OpenFOAM-based framework was used to assess the effects of CPB pump flow rate (3-5 LPM), hemodiluted blood viscosity (1.5–3.5 cP), and embolus size (0.5–2.5 mm) on embolic distribution across major abdominal branches, including the renal, hepatic, splenic, mesenteric, and iliac arteries. Key results showed that lower blood viscosity (1.5 cP) and higher flow rate (5 LPM) significantly affected embolic transport, both individually and combinedly. Under these combined conditions, renal artery emboli transport increased from 17% to 27%, and hepatic artery transport rose from 7.1% to 10.7%. Also, larger emboli (2.5 mm) consistently exhibited greater deviation from central flow and had higher side-branch entry, resulting in increased escape rates of 14% and 26% for renal and hepatic branches, respectively. These tendencies correlated well with clinical observations of post-CPB acute kidney and liver injury. This study presents the first CFD-based quantitative analysis of embolic transport to abdominal organs, revealing critical emboli pathways previously overlooked in both clinical and computational studies. The findings highlight the need for optimized CPB perfusion strategies to minimize embolic burden and enhance intraoperative protection of abdominal organs during cardiac surgery.

## 1. Introduction

Cardiopulmonary bypass (CPB) is performed globally at an increasing rate, with over 500,000 procedures conducted annually in the United States alone (Tsao et al. 2023). While CPB has revolutionized cardiac surgery, it is associated with significant systemic effects as blood is artificially circulated through vital organs during the procedure. The physiological stress of CPB is aggravated by interventions such as hemodilution, pump flow rate variations, and non-pulsatile flows, which can significantly impact perfusion dynamics across all organ systems. Despite these risks, the specific effects of CPB pump operating conditions on abdominal artery thrombosis and visceral organ function remain poorly understood, necessitating further investigation to optimize CPB management strategies and mitigate complications.

Advancements in computational fluid dynamics (CFD) have enabled complex numerical simulations of cardiovascular hemodynamics, facilitating detailed analyses of flow behavior and pathological conditions. When integrated with Lagrangian particle tracking (LPT), CFD allows precise tracking of emboli trajectories, enabling the assessment of embolism risk and infarction potential. Prior studies have employed this capability to investigate emboli transport in the thoracic aorta (Arefin and Good 2024; Khamooshi et al. 2024) and the Circle of Willis (**2751**; Mukherjee, Padilla, and Shadden 2016), particularly in the context of acute ischemic stroke. More recently, this approach has been applied to the ECMO extracorporeal system to examine emboli dynamics in the full-length aorta (Khamooshi et al. 2024). To date, CFD-based hemodynamic studies of abdominal aortic flow remain scarce (Al-Rawi and Al-Jumaily 2016), with a notable lack of research on emboli transport through the renal, hepatic, splenic, and mesenteric arteries. Despite growing clinical evidence linking CPB to complications in the kidneys, liver, spleen, pancreas, gallbladder, and gastrointestinal organs, no studies have explored the influence of CPB pump operating conditions on these critical visceral organs.

Emboli traveling through the descending aorta can lodge within the renal arteries (RAs), obstructing blood flow and potentially leading to acute kidney injury (AKI) with severe consequences (Granata et al. 2012; O’Neal, Shaw, and Billings 2016). This obstruction, characterized by diminished renal perfusion, poses a significant risk to kidney function, as complete occlusion can result in irreversible renal failure. AKI occurs in 18.2% to 30% of patients undergoing CPB (Rosner and Okusa 2006; Haase et al. 2012; Krawczeski et al. 2011) and is a key predictor of post-cardiac surgery morbidity and mortality (Hobson et al. 2009; Bahar et al. 2005). CPB surgery is one of the two most common causes of AKI (Haase-Fielitz et al. 2007), which is associated with prolonged hospital stays, poor clinical outcomes, and an eightfold increase in hospital mortality (Long, Jenkins, and Griffith 2015; Pickering, James, and Palmer 2015; Luckraz et al. 2005). While cerebral and renal embolization have been studied, hepatic embolization remains relatively underexplored despite its critical role in postoperative liver dysfunction (Di Tomasso, Monaco, and Landoni 2015). Hemodynamic alterations during CPB, including non-pulsatile perfusion and hemodilution, increase the risk of embolic entrapment within the hepatic microcirculation (Di Tomasso, Monaco, and Landoni 2015). The incidence of hepatic failure after cardiac surgery is approximately 4%, with up to 10% of CPB patients experiencing some degree of liver injury (Edmunds 2004; Michalopoulos, Alivizatos, and Geroulanos 1997). A separate study reported a 14.4% incidence of post-CPB liver dysfunction in patients without other intra-abdominal complications (Ohri et al. 1991). Hepatic failure accounts for approximately 4% of visceral complications but carries a staggering 74% mortality rate (Sakorafas and Tsiotos 1999).

Despite the spleen’s rich blood supply, reports of splenic embolism remain relatively scarce (Hazanov et al. 2006). However, embolic events are increasingly recognized as a major cause of splenic infarction in post-cardiac surgery patients. A clinical autopsy study of post-cardiac surgery cases found thromboembolic events responsible for 67% of splenic infarctions (O’Keefe Jr et al., n.d.), highlighting embolism as a major contributing factor. Another study reported that one-third of splenic infarction cases were embolic in origin (Jaroch, Broughan, and Hermann 1986). Mortality rates associated with splenic infarction vary widely, ranging from 5% to 20%, depending on patient demographics and underlying conditions (Hazanov et al. 2006).

Other abdominal visceral organs have also been shown to be severely impacted by ischemic complications that can occur during cardiac surgery, including the gastrointestinal system (Ohri and Velissaris 2006) and the pancreas (Lefor et al. 1992). While relatively rare, reported incidence rates of CPB-induced gastrointestinal dysfunction range from 0.3-5.5%, but with reported mortality rates of approximately 32% (Rodriguez et al. 2010; McSweeney et al. 2004). Similarly, pancreatic complications following CPB are uncommon (0.44% of cases) but carry a high mortality risk (44% (Lefor et al. 1992)), with pancreatic ischemia identified as a major contributing factor. While enough clinical data exists on CPB-related complications in abdominal visceral organs, the associated embolism risk remains largely unexplored and unquantified. This study aims to bridge this gap by utilizing CFD with LPT to model emboli transport through key abdominal arteries, including the renal, hepatic, splenic, mesenteric, and common iliac. This study will provide a detailed assessment of embolic transport dynamics in the abdominal circulation by evaluating the influence of CPB operating conditions on perfusion and embolism risk, as well as mapping emboli trajectories and lodgment patterns in the kidneys, liver, intestines, spleen, gallbladder, and pancreas. These findings will improve our understanding of CPB-induced embolization, guiding risk mitigation strategies and refining perfusion management during CPB.

## 2. Methods

### 2.1 CFD Model Development

A patient-specific CFD model of the aorta was developed to simulate embolic transport under CPB conditions. The aortic geometry, encompassing both the thoracic and abdominal regions, was obtained from the Vascular Model Repository (Wilson, Ortiz, and Johnson 2013) and represents a 21-year-old female with no preexisting cardiovascular disease (Fig. 1). To enable emboli injection, a cylindrical CPB outlet cannula, with dimensions (20 French, 6.67 mm diameter) in accordance with established clinical standards (Hospital 2004; Health 2019), was incorporated into the model using Blender, an open-source software for 3D geometry processing and refinement. Blender was further utilized to eliminate extraneous anatomical features and define the inlet and outlet boundary patches of the aortic model for subsequent CFD simulations (Fig. 1). In total, the model includes one inlet (CPB outlet cannula) and ten outlets (brachiocephalic artery (BCA), left common carotid artery (LCCA), left subclavian artery (LSCA), left and right RAs, common hepatic artery (HA), splenic artery (SA), superior mesenteric artery (SMA), and left and right iliac arteries (IAs). Note that during CPB, the ascending aorta is cross-clamped, therefore, it is modeled as a closed wall with no flow entering the model via the left ventricle.

**Figure 1.**
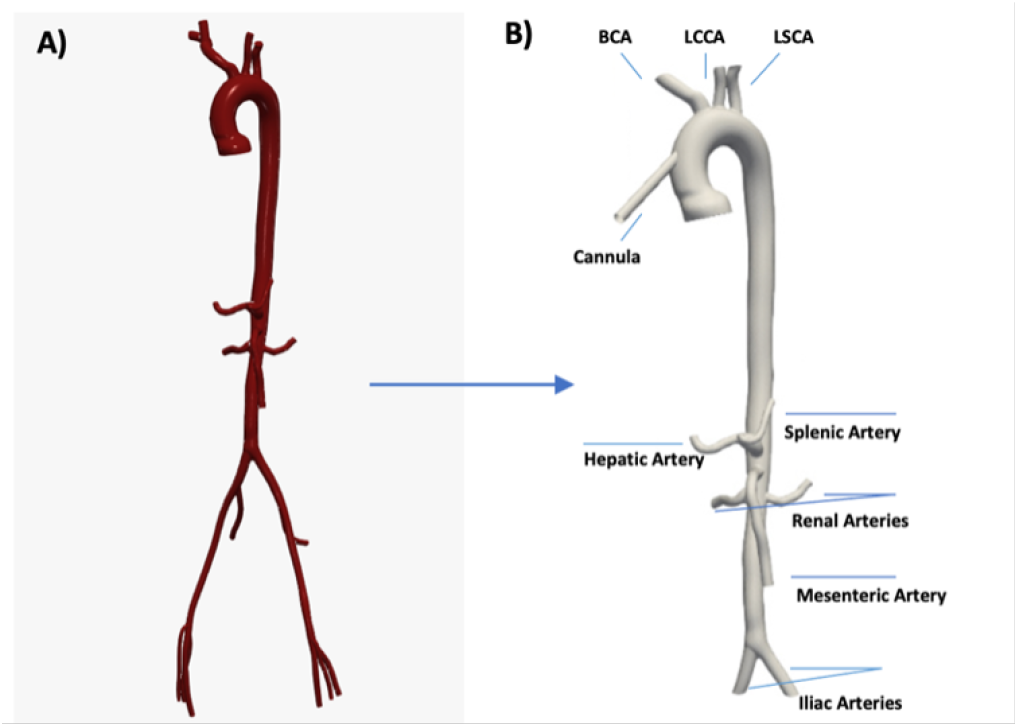
A) Patient-specific aorta model from the Vascular Model Repository. B) Developed CAD model with an anastomosed CPB outlet cannula.

The simulations were conducted using OpenFOAM, an open-source C++ computational continuum mechanics library. The Reynolds-averaged Navier-Stokes (RANS) equations were solved under continuous CPB inlet flow using OpenFOAM’s ‘pisoFoam’ solver, which employs the finite volume method and the Pressure-Implicit with Splitting of Operators (PISO) algorithm to compute the velocity and pressure fields. A generalized geometric/algebraic multi-grid (GAMG) solver was employed for pressure, while a preconditioned bi-conjugate gradient (PBICG) solver was applied to velocity calculations. To ensure numerical stability and accuracy, a dynamic time-stepping strategy was implemented to maintain a Courant number below 1, with all residuals converging to 10 at each time step.

To accurately resolve turbulent flow characteristics at the CPB cannula outlet, where peak Reynolds numbers reach approximately 7,195 and 11,990 for 3 LPM and 5 LPM CPB flow rates, respectively, the k-omega shear-stress transport (k-omega SST) turbulence model (Menter 1994) was employed. This turbulence model was preferred for its superior handling of adverse pressure gradients, separated flows, and transitional flow regimes (Menter, Kuntz, and Langtry 2003). It combines the strengths of the k-omega model near solid boundaries with the k-varepsilon model in the free-stream region, improving predictions in adverse pressure gradients and separated flows. This approach (RANS with a k-omega SST turbulence model) was employed in our prior work investigating CPB flows in the aortic arch (Arefin and Good 2024) and shown to be equivalent in predictions of emboli transport compared to the more computationally intensive large eddy simulation (LES) approach with a Smagorinsky subgrid-scale model.

A systematic mesh sensitivity analysis was undertaken to ensure numerical independence from mesh resolution. Unstructured grids of varying refinement levels were generated using OpenFOAM’s ‘snappyHexMesh’ utility, consisting of coarse (995,000 cells), medium (2,140,000 cells), and fine (2,997,000 cells) resolutions. Each mesh was predominantly comprised of hexahedral interior cells, with three additional near-wall layers incorporated to enhance boundary layer resolution. To assess how mesh refinement influenced emboli transport, two CPB flow cases were simulated and compared across each mesh. Case A featured the highest viscosity fluid (3.5 cP) at a pump flow rate of 3 LPM with emboli of 2.5 mm diameter, while Case B utilized the lowest viscosity fluid (1.5 cP), keeping all other parameters identical. The resulting simulations were analyzed to evaluate the influence of mesh resolution on embolic transport dynamics.

### 2.2 CFD Boundary Conditions

CFD boundary conditions were applied to closely replicate the physiological conditions observed in human cardiovascular circulation. Pressure boundary conditions were applied at the outlets of the three aortic arch branches and the seven abdominal aorta branches to ensure accurate flow distributions, governed by Eq. 1:

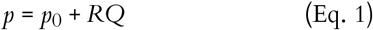

where p is the outlet pressure, p0 is the mean arterial pressure, R is a resistance constant, and Q denotes the flow rate through the outlet boundary. An iterative simulation approach was employed to determine the appropriate resistance coefficient (R) for each outlet, ensuring accurate physiological flow distributions across three different blood viscosities (1.5, 2.5, and 3.5 cP) and two flow rate conditions (3 and 5 LPM). This resulted in n=6 distinct sets of resistance coefficients corresponding to validated flow distributions of 15.7% through the BCA, 7.8% through the LCCA, and 7.8% through the LSCA (Benim et al. 2011). In the abdominal branches, flow distributions were 20% combined through the RAs (Castenfors 1977), 6.9% through the HA (Carlisle et al. 1992), 5% through the SA (Warkentin 2018), 12% through the SMA supplying the intestines (Norryd et al. 1975), and the remainder exiting through the IAs. Additionally, zero-gradient velocity boundary conditions were imposed at all outlets to maintain consistency in outflow behavior, while a no-slip velocity boundary condition was enforced on all vessel walls.

### 2.3 Lagrangian Particle Tracking

LPT was employed to analyze the distribution of emboli through the aorta. LPT is a volumetric flow measurement technique that enables the tracking of individual tracer particles over extended time periods even in highly turbulent flow regimes (Yeoh and Tu 2019). In this study, embolic transport was simulated using the validated LPT solver ‘particleFoam’ within OpenFOAM. The Lagrangian directory ‘cloudProperties’ in OpenFOAM facilitates particle injection using various built-in models and tracks their movement based on specified forces. Here, each parcel was modeled as an individual embolus, with a manual injection model employed to precisely control the size and initial position of each particle within the fluid domain.

To ensure statistical significance in embolic distribution analysis, 1000 particles were injected into the aortic model through the CPB cannula inlet in each simulation. The particles were introduced at the same velocity as the inflowing blood from the CPB circuit, while particle collisions were disabled, assuming that only a single embolus would be injected from the CPB circuit into the aorta at a time. Particles were modeled to rebound upon contact with the wall using the ‘patchInteractionModel’ within ‘cloudProperties’. The physiological characteristics of the arterial wall were represented through its material properties defined by a Young’s modulus of 4.66e6 and a Poisson’s ratio of 0.45 (Giannakoulas et al. 2005), implemented using the ‘wallModel’ tool in OpenFOAM.

Initial flow simulations were performed for 2.5 seconds to establish baseline flow conditions throughout the aorta and ensure that aortic branch outlet flows reached a steady state before initiating embolus injection. Embolic transport simulations using LPT were performed for an additional 40 seconds to ensure complete passage of all injected emboli through one of the thoracic or abdominal aortic outlet boundaries. Simulation results were post-processed and visualized using ParaView (Fig. 2). During phase (i), emboli were introduced into the aortic model through the CPB outlet cannula. In phase (ii), their progression toward the outlets can be observed, and by phase (iv), the majority have exited the fluid domain.

**Figure 2.**
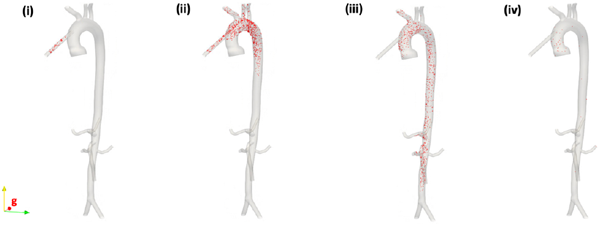
Emboli transport through the aorta over time, shown for one representative simulation case.

Particle motion was modeled using a Lagrangian framework, in which the trajectory of each embolus was computed by solving a system of ordinary differential equations (ODEs) along its path. A Euler first-order integration scheme was employed to update particle position and velocity over time. This approach enabled the calculation of both translational and rotational kinematics. It accounted for the dominant physical forces acting on the particles, with particular emphasis on sphere drag (Eq. 6), gravity (Eq. 8), and local pressure gradients (Eq. 9), ensuring physiologically relevant simulation of embolic transport dynamics. The translational motion of a particle was governed by Eq. 2:

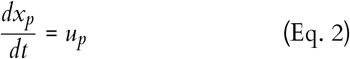

where *x*_*p*_ is the particle’s position and *u*_*p*_ is its linear velocity. Its rotational motion was governed by Eq. 3:

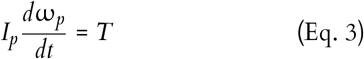

where *I*_*p*_ is the particle’s moment of inertia, ω_*p*_ is its angular velocity, and *T* is the net torque. For the rigid spherical emboli in our simulation, *I*_*p*_ is calculated by Eq. 4:

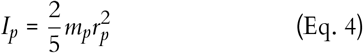

where *m*_*p*_ is the particle’s mass and *r*_*p*_ is its radius. The net force acting on each particle was computed using Newton’s second law, expressed as Eq. 5:

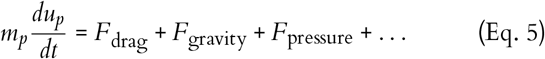

Here, the terms on the right side of the equation represent the dominant forces considered. The drag force (*F*_drag_), a major contributor to particle motion, was modeled using a mass-based formulation suitable for LPT solvers (Eq. 6):

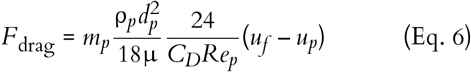

Here (*u*_*f*_ – *u*_*p*_) represents the relative velocity between the fluid and the particle. The drag coefficient, *C*_*D*_, in Eq. 6 varies with the particle Reynolds number *Re*_*p*_. For *Re*_*p*_ < 1000, the Schiller–Naumann correlation was applied to capture viscous and inertial effects (Schiller 1933) (Eq. 7a), while for *Re*_*p*_ *≥* 1000, a constant value of *C*_*D*_ = 0.44 was adopted to approximate the drag force in the turbulent regime, based on empirical data (Clift, Grace, and Weber 2005):

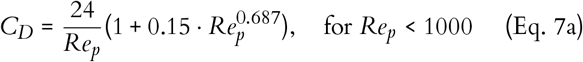

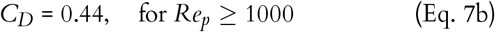

The gravitational force (*F*_gravity_), acting on the particle is given by Eq. 8, while the pressure gradient force (*F*_pressure_), which arises due to spatial pressure variations in the fluid, is expressed as Eq. 9:

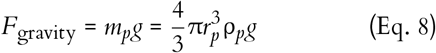

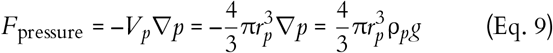

These equations form the mathematical basis of our LPT approach, enabling the computation of embolic trajectories through direct integration of force balances along individual particle paths, thereby allowing for the accurate simulation of embolic transport within anatomically realistic thoracic and abdominal aortic flow fields.

### 2.4 CPB Parameters

To account for the range of clinically relevant hemodiluted blood viscosities that can occur during CPB, three different fluid viscosities were considered: 1.5, 2.5, and 3.5 cP. The CPB pump flow rate, measured in liters per minute (LPM), is clinically determined as the product of a patient’s body surface area (BSA) and a standard cardiac index factor ranging between 2.2 and 2.4 (Jw et al., n.d.; Starr 1959). The BSA is calculated using the Du Bois formula (Eq. 10), which accounts for the patient’s height and weight under the assumption that each square meter of body surface maintains a uniform metabolic rate (Bois 1989):

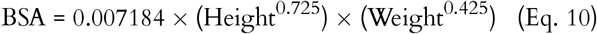

The Du Bois equation was utilized to determine representative BSA values, enabling the selection of pump flow rates for two patient groups at opposite ends of the BSA spectrum. Consequently, flow rates of 3 and 5 LPM were chosen to represent patients with lower and higher BSAs, respectively. These values were applied across the computational simulations to analyze the impact of CPB flow rate on emboli transport.

### 2.5 Emboli Parameters

To investigate the influence of embolus size on transport behavior, emboli of varying diameters were introduced in the simulations. Prior studies have demonstrated that emboli smaller than 0.2 mm tend to follow the fluid streamlines proportionally without significant deviation (Chung et al. 2010). Based on this finding, microemboli were excluded from this study. Also, clinical observations indicate that emboli found in the aorta can range from 0.3 to 2.9 mm in diameter (Barbut et al. 1997). Therefore, to ensure physiologically relevant conditions, three distinct emboli sizes were selected for investigation: 0.5, 1.5, and 2.5 mm. By incorporating emboli with different physical characteristics, the study aimed to comprehensively evaluate embolic behavior under varying CPB flow and viscosity conditions.

### 2.6 Prior Experimental Validation Supporting Current CFD Framework

In our prior study on CPB emboli transport using the same OpenFOAM solvers (‘pisoFoam’ and ‘particleFoam’), model setup, LPT approach, and boundary conditions, we experimentally validated the computational results using a mock circulatory flow loop with a 3D-printed patient-specific silicone aorta model (Arefin and Good 2024). The in vitro trials employed over 174 experiments per case, as determined by a sample size calculation (90% confidence level, 5% margin of error), to ensure statistical power in emboli exit distribution. The experimental validation confirmed the accuracy and reliability of our CFD simulations. Numerically predicted emboli distributions closely matched measured values across varying flow rates, emboli sizes, and blood viscosities, with deviations < 6.2% in all cases. Given that the present study employs identical numerical methods, boundary conditions, CPB flow parameters, and particle properties, apart from minor differences in anatomical details and the addition of the abdominal aortic region, we confidently consider the current CFD-LPT results to be built upon a previously experimentally validated modeling framework.

## 3. Results

### 3.1 Grid Convergence

Emboli exit through the aortic arch branches and RAs for two CPB cases were compared to determine the influence of grid refinement on resultant predictions of emboli transport. Both cases involved 2.5 mm emboli and 3.5 cP blood. Case A featured a lower pump flow rate of 3 LPM, while Case B employed a higher flow rate of 5 LPM. In Case A, emboli exiting through the aortic arch branches exhibited a 9.1% difference between the coarse and medium meshes, which reduced to 4.9% between the medium and fine meshes (Fig. 3). Similarly, for emboli exiting through the RAs, the corresponding differences were 15.1% and 3.6% between the coarse-medium and medium-fine meshes, respectively. In Case B, emboli exiting through the aortic arch branches showed a 14.8% difference between the coarse and medium meshes, decreasing to 5.3% between the medium and fine meshes. For the RAs, the differences were 24.9% and 4.9% between coarse-medium and medium-fine meshes, respectively.

**Figure 3.**
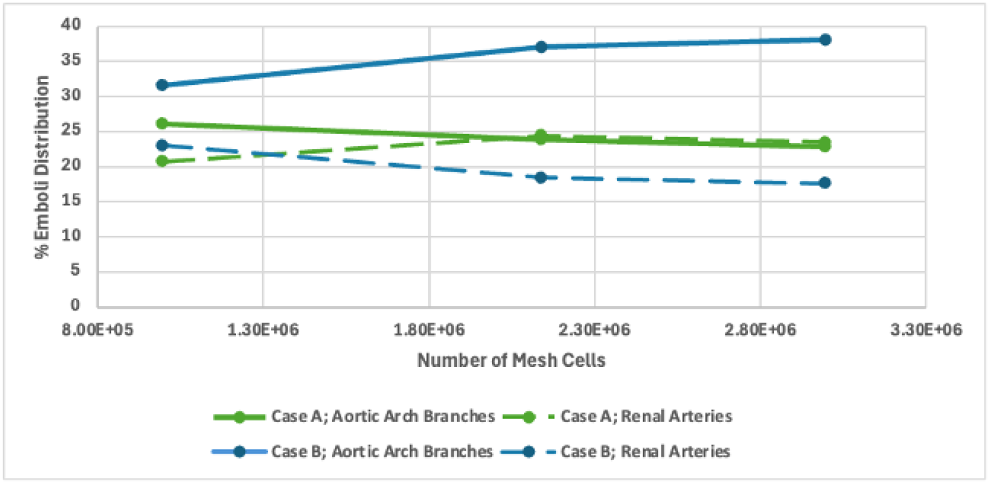
Effect of mesh refinement on emboli transport into the aortic arch branches and renal arteries

To quantify the numerical uncertainty introduced by discretization, the Grid Convergence Index (GCI) was calculated using generalized Richardson extrapolation (Richardson and Gaunt 1927). In accordance with Roache’s methodology for unstructured meshes, a factor of safety of 1.25 was used for unstructured meshes to provide an upper limit on the relative discretization error (Roache 1994). The GCI values for Case A were 11.9% and 6.4% for the medium and fine meshes in the aortic arch branches, and 5.6% and 1.4% in the RAs, respectively. For Case B, the GCIs were 9.5% and 3.4% for the aortic arch branches, and 7.1% and 1.5% for the RAs, respectively. The consistently decreasing solution differences between medium and fine meshes, along with low GCI values, indicated asymptotic convergence. Therefore, the medium-mesh (2.1 million cells) was considered sufficiently refined and used for all subsequent CFD analyses.

### 3.2 Effect of CPB Pump Flow Rate

Simulations were performed at pump flow rates of 3 and 5 LPM, incorporating the extremes of body surface area (BSA), to assess emboli distribution. Fig. 3 illustrates the average percentage of emboli exiting through the aortic arch branches (potentially causing a stroke), renal, hepatic, splenic, and mesenteric arteries (potentially leading to abdominal organ injury) during CPB.

The results represent a combination of all three emboli sizes and the three hemodiluted blood viscosity conditions. The RAs showed the most pronounced increase in embolic transport from 3 to 5 LPM, rising by 29%, while the other arterial branches demonstrated more modest increases under the same conditions. The SAs experienced nearly identical embolic transport under both CBP flow rates.

### 3.3 Effect of Hemodiluted Blood Viscosity

As the influence of hemodilution was assessed, simulations were performed at blood viscosities of 3.5, 2.5, and 1.5 cP. As blood viscosity decreased with increased hemodilution, a general rise in the percentage of emboli exiting the arteries was observed (Fig. 4). The most pronounced effect was observed in the RAs and HA, where emboli transport increased by 18% and 35%, respectively, when blood viscosity was reduced to 1.5 cP.

**Figure 4.**
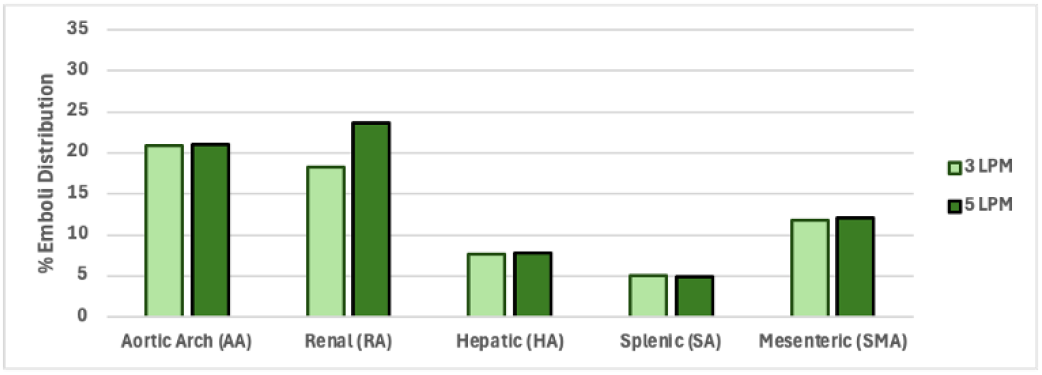
Emboli transport in the aortic branches for varying CPB pump flow rates.

**Figure 5.**
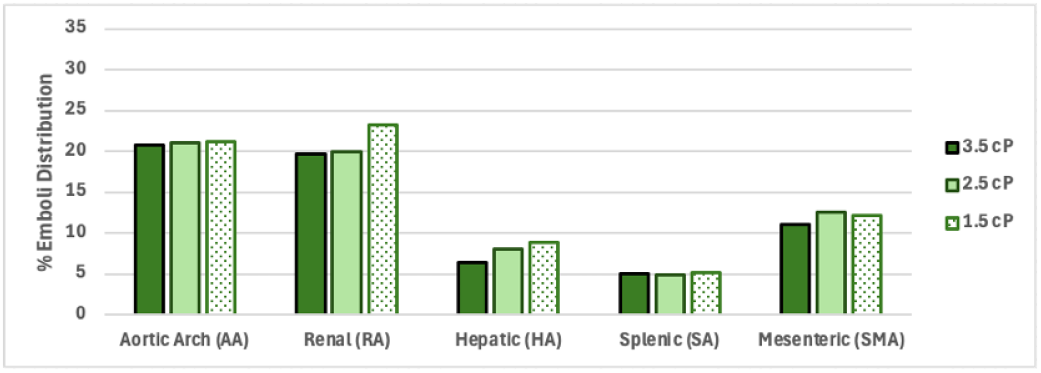
Emboli transport in the aortic branches for varying hemodiluted blood viscosities.

**Figure 6.**
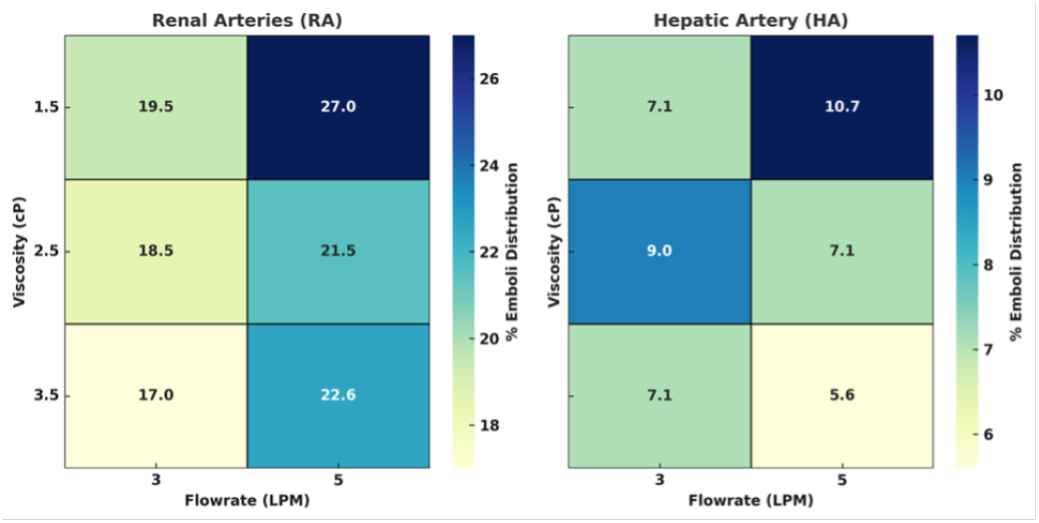
Heatmaps illustrating the combined effect of varying CPB flow rate and blood viscosity on particle exit distribution through the RAs and HA.

The heatmap in Fig. 6 highlights the significant variability in emboli transport across different CPB flow rates and blood viscosity conditions. Among all aortic branches, Ras consistently received higher emboli loads, while distribution in both the RAs and the HA peaked under conditions of lowest viscosity (1.5 cP) and highest flow rate (5 LPM). In the RAs, emboli transport reached a maximum average of 27% under these conditions and decreased with increasing blood viscosity, dropping to 17% at 3.5 cP and 3 LPM. At a fixed viscosity, increasing the flow rate from 3 to 5 LPM consistently elevated emboli transport in the RAs, with the most substantial rise observed at 1.5 cP, where emboli exit increased by 35.5%. The HA exhibited a similar trend, though to a lesser extent. At 1.5 cP, emboli transport increased from 7.1% at 3 LPM to 10.7% at 5 LPM, while changes at higher viscosities remained minimal. Overall, the heatmaps illustrate that both CPB flow rate and blood viscosity independently and collectively influence emboli transport, with viscosity showing a stronger inverse correlation to emboli exit, especially within renal circulation.

### 3.4 Effect of Emboli Size

The relationship between emboli size and pump flow rate is illustrated in Fig. 7, which presents the total percentage of emboli that exited through the aortic branches (except the IAs traveling to the legs), aggregated across all hemodiluted blood viscosity conditions. The results indicate that larger emboli are more likely to exit through the thoracic and abdominal aortic branches, as opposed to exiting through the IAs, at both flow rates. Specifically, emboli transport to the branches increased progressively with emboli size, with an overall 25% to 44% rise observed when comparing emboli sizes of 0.5 and 2.5 mm.

**Figure 7.**
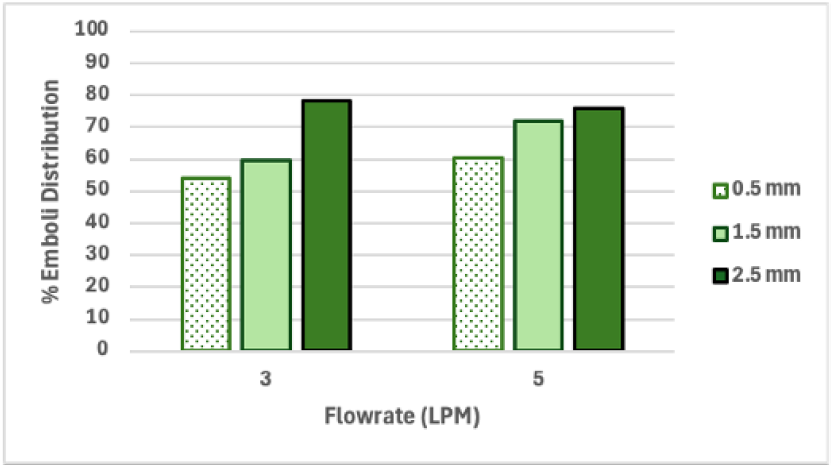
Emboli transport in the aortic branches at two CPB flowrates (3 and 5 LPM) across varying emboli sizes.

The influence of large emboli (1.5 and 2.5mm) transport through the aortic and abdominal branches was further differentiated in Fig. 8. The results indicate a positive correlation between embolus size and transport rates, with larger emboli demonstrating greater exit percentages into all of the aortic branches. Notably, emboli transport through the RAs increased by 14% for 2.5 mm emboli compared to 1.5 mm emboli. A detailed explanation of size-dependent embolic dynamics is provided in Section 4.

**Figure 8.**
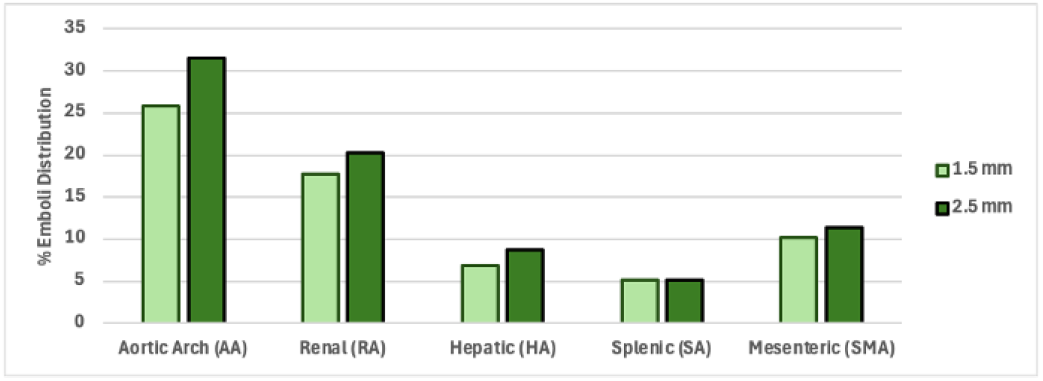
Large emboli transport in the aortic branches

## 4. Discussion

The present study provides critical understanding of the complex dynamics of emboli transport within the thoracic and abdominal aorta. Utilizing previously validated CFD models, we investigated the influence of key CPB parameters including blood viscosity, pump flow rate, and embolus size, on embolius distribution within all major thoracic and abdominal aortic branches. The in-silico models replicated realistic CPB conditions and allowed for detailed predictions of emboli trajectories, offering valuable implications for minimizing organ-specific injury risks such as AIS and AKI.

Our findings reveal that embolus size is a dominant factor in determining exit behavior through the arterial branches. As emboli size increases, they exhibit a greater tendency to exit from the aortic arch branches and descending aorta branches, regardless of flow rate (Fig. 6). The difference in escape rates between the smallest (0.5 mm) and largest (2.5 mm) emboli ranged from 25% to 44%, with larger emboli exhibiting significantly higher exit probabilities. This behavior can be explained by the Stokes number (St, Eq. 11), which characterizes a particle’s ability to follow fluid streamlines:.

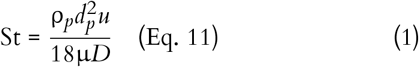

where the particle diameter, *d*_*p*_, in the numerator is squared and has a greater impact on St compared to the particle’s density, ρ_*p*_, and the fluid’s viscosity, µ. Smaller emboli, with lower St, experience greater drag and reduced inertia, causing them to remain tightly coupled with the central flow streamlines and reducing their chances of exiting into side branches. In contrast, larger emboli deviate more readily from these streamlines due to their higher inertia, increasing their propensity to migrate toward the aortic branch outlets. The effect of St on overall emboli transport dynamics is shown in Figure 9.

**Figure 9.**
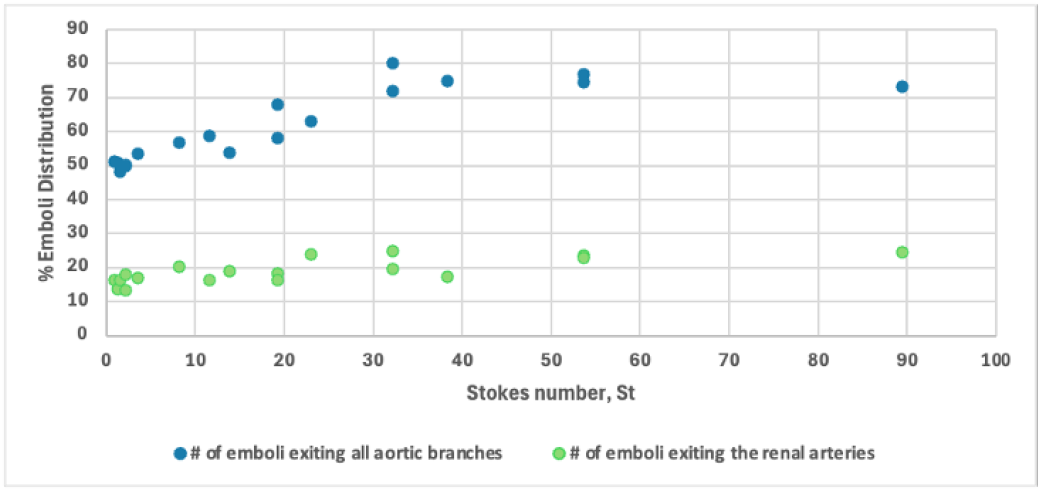
Emboli transport as a function of the Stokes number (St).

Further, blood viscosity itself emerged as a key determinant of emboli distribution. Our analysis showed that reduced blood viscosity, resulting from hemodilution that occurs during CPB, leads to an increase in emboli escape through both the thoracic and abdominal aortic branches (Fig. 5). The RAs and HA showed the greatest rise in emboli transport, by 18% and 35%, respectively. Hemodiluted blood, characterized by decreased red blood cell concentration, results in reduced shear stresses along vessel walls and facilitates emboli migration toward these slower-flowing near-wall regions of the vasculature, thereby increasing the probability of emboli exiting through the branching arteries. These results highlight the profound influence of blood rheology on emboli transport dynamics during CPB.

Additionally, increasing the pump flow rate from 3 LPM to 5 LPM resulted in greater emboli exit rates through the aortic branches, with the most pronounced effect observed in the RAs, where emboli transport increased by 29% under the higher flow condition (Fig. 4). This can potentially be attributed to increased turbulence generated by the cannula outflow entering the ascending aorta, where the Reynolds number rose from 3,410 to 5,680 for normal blood (3.5 cP), and from 7,195 to 11,990 for the highly diluted blood (1.5 cP). Increased turbulence disrupts laminar flow patterns and promotes emboli deviation from their initial streamlines. Such turbulence-induced deviation reinforces the importance of carefully regulated pump flow rate to minimize embolic complications.

These findings are consistent with our previous study conducted on a different patient-specific model limited to the thoracic aorta (Arefin and Good 2024), where higher emboli exit through the aortic branches was similarly observed under increased pump flow rate and reduced hemodiluted blood viscosity conditions. Also, our simulation results align with clinical observations of organ injury during CPB, particularly in the kidneys, liver, and spleen. The increased emboli exit rates into the RAs at higher pump flow conditions (up to 29%) observed in this study are consistent with the high incidence of AKI reported in 18.2–30% of CPB patients (Rosner and Okusa 2006; Haase et al. 2012; Krawczeski et al. 2011), where embolic obstruction is a known contributor to impaired renal perfusion and subsequent renal failure (Granata et al. 2012). Similarly, elevated embolic delivery to the HA and SA under low-viscosity and high-flow conditions correlates with the reported 4–14.4% incidence of liver dysfunction and 67% embolic origin of splenic infarctions following cardiac surgery (Michalopoulos, Alivizatos, and Geroulanos 1997; Edmunds 2004; Ohri et al. 1991; O’Keefe Jr et al., n.d.). These correlations support the hypothesis that emboli generated during CPB may lodge in critical abdominal organs, contributing to postoperative complications. By mapping emboli trajectories into these arterial branches, our study provides a quantitative foundation for understanding the mechanism of CPB-induced visceral organ injury, which has previously been clinically recognized but biomechanically underexplored.

This research has certain limitations to be acknowledged. First, the emboli were modeled as rigid, spherical particles, which do not fully capture the range of morphologies and mechanical properties observed in vivo. Real emboli may have irregular shapes and viscoelastic characteristics, which can influence their interactions with vascular walls and their transport through the complex vasculature. Additionally, our simulations assumed steady-state flow conditions, which, while appropriate for CPB scenarios with a clamped ascending aorta, are not fully reflective of the pulsatile flow patterns present under normal physiological conditions. Future research incorporating pulsatile flow inlet boundary conditions, vessel wall compliance, and more physiologically accurate embolus models could improve the predictive power of these simulations.

## 5. Conclusion

This study presents the first comprehensive patient-specific computational analysis of emboli transport into the descending thoracic and abdominal aorta during CPB. Using a previously validated framework integrating computational fluid dynamics (CFD) and Lagrangian particle tracking (LPT), we simulated embolic trajectories across all aortic branches under six clinically relevant CPB conditions, resulting in a total of 18 simulations, tracking 1000 emboli per case.

Key results demonstrate that emboli transport is significantly affected by both flow and rheological parameters, with lower blood viscosity (1.5 cP) and higher flow rate (5 LPM) significantly increasing embolic migration into the abdominal arterial branches. Specifically, renal artery emboli transport increased from 17% to 27%, while hepatic artery emboli exit rose from 7.1% to 10.7% under these conditions. Larger emboli (2.5 mm) were also more likely to deviate from central flow paths and enter side branches, with overall embolic exit increasing by 25–44% as emboli diameter increased. These trends were supported by Stokes number-based analysis, providing a mechanistic perception into the role of embolus inertia and fluid-particle coupling.

Based on these findings, we recommend that CPB pump flow rates be managed judiciously and not routinely set at the upper limits dictated by patient BSA. Likewise, excessive hemodilution should also be minimized when feasible, to reduce embolic dispersion. However, strict adherence to these guidelines may not always be clinically feasible. In certain patients, higher flow rates may be necessary to maintain adequate tissue perfusion, particularly in cases of elevated metabolic demand, low hematocrit, or impaired oxygen delivery. Similarly, hemodilution is sometimes indispensable to reduce blood viscosity and improve circuit flow.

These recommendations aim to reduce embolic transport into critical arterial branches and mitigate the risk of acute ischemic stroke (AIS), acute kidney injury (AKI), hepatic dysfunction, and other abdominal arterial complications. Beyond these clinical implications, this study provides the first comprehensive analysis of emboli transport into the descending aorta and major abdominal arterial branches under CPB conditions, a region largely overlooked in previous CFD studies, which have primarily focused on the aortic arch and cerebral circulation. By modeling emboli trajectories into the renal, hepatic, splenic, and mesenteric arteries, this work identifies previously unquantified embolic pathways that may contribute to visceral organ injury. The impact of this study extends beyond computational modeling, offering actionable guidance for surgical planning and intraoperative management. These findings bridge a critical gap between clinical observations of postoperative organ injuries and their underlying flow-driven mechanisms, providing a strong foundation for CPB risk assessment and the optimization of perfusion strategies to protect both cerebral and abdominal organs during cardiac surgery.

## Data Availability

All data produced in the present work are contained in the manuscript

## Author Contributions

Conceptualization: N.A; B.G. Methodology: N.A; B.G. Data curation: N.A. Data Visualization: N.A. Resources: B.G. Writing - original draft: N.A. Writing - review and editing: N.A; B.G. Software: N.A. Formal Analysis: N.A. Supervision: B.G. Project Administration: B.G. All authors approved the final submitted draft.

## Ethical Standards

The research meets all ethical guidelines, including adherence to the legal requirements of the study country.

